# Dysregulation of tryptophan catabolism at the host-skin microbiota interface in Hidradenitis Suppurativa

**DOI:** 10.1101/2020.08.25.20181438

**Authors:** Laure Guenin-Macé, Jean-David Morel, Jean-Marc Doisne, Angèle Schiavo, Lysiane Boulet, Véronique Mayau, Pedro Goncalves, Sabine Duchatelet, Alain Hovnanian, Vincent Bondet, Darragh Duffy, Marie-Noëlle Ungeheuer, Maïa Delage, Aude Nassif, James P Di Santo, Caroline Demangel

**Affiliations:** Immunobiology of Infection Unit, Institut Pasteur, INSERM U1221, Paris, France; Université Paris 7, Sorbonne Paris Cité, Paris, France; ENS de Lyon, Lyon, France; Innate Immunity Unit, Institut Pasteur, INSERM U1223, Paris, France; Laboratoire de Biochimie Hormonale et Nutritionnelle, CHU Grenoble-Alpes, La Tronche, France; Université de Paris, Imagine Institute, Laboratory of Genetic Skin Diseases, INSERM UMR 1163, Paris, France; Department of Genetics, AP-HP, Hôpital Necker-Enfants Malades, Paris, France; Immunobiology of Dendritic Cells, Institut Pasteur, INSERM U1223, Paris, France; ICAReB platform, Institut Pasteur, Paris, France; Centre Médical, Institut Pasteur, Paris, France

**Author notes:** These authors contributed equally (Laure Guenin-Macé, Jean-David Morel). **Corresponding author**: Caroline Demangel, Institut Pasteur, 25 Rue du Dr Roux, 75724 Paris Cedex 15, France.

## Abstract

Hidradenitis Suppurativa (HS) is a chronic skin disorder of unknown etiology that manifests as recurrent, painful lesions. Cutaneous dysbiosis and unresolved inflammation are hallmarks of active HS, but their origin and interplay remain unclear. Our metabolomic profiling of HS skin revealed an abnormal induction of the kynurenine pathway (KP) of tryptophan catabolism in dermal fibroblasts correlating with the release of KP-inducing cytokines by inflammatory cell infiltrates. Notably, over-activation of the KP in lesional skin was associated with local and systemic depletion in tryptophan. Yet the skin microbiota normally degrades host tryptophan into indoles regulating tissue inflammation via engagement of the Aryl Hydrocarbon Receptor (AHR). In HS skin lesions, we detected contextual defects in AHR activation coinciding with impaired production of bacteria-derived AHR agonists and decreased incidence of AHR ligand-producing bacteria in the resident flora. Dysregulation of tryptophan catabolism at the skin-microbiota interface thus provides a mechanism linking the immunological and microbiological features of HS lesions. In addition to revealing metabolic alterations in HS patients, our study suggests that correcting AHR signaling would help restore immune homeostasis in HS skin.

## Introduction

Hidradenitis Suppurativa (HS), also known as Verneuil’s disease and *acne inversa*, is an inflammatory disease of the pilosebaceous follicle typically starting in the second decade of life. The average prevalence of HS is 1% with a 3-fold greater incidence in women (1). In most patients (68%) HS manifests as recurrent subcutaneous nodules or abscesses (Stage I lesions in Hurley’s severity classification). Some patients develop more severe forms of the disease, with fistulas (Hurley II, 28%) or multiple interconnecting lesions (Hurley III, 4%). In addition to causing intense pain, HS is commonly associated with depressive symptoms and anxiety (2). There is no standard treatment or definitive cure for HS, but immunomodulatory drugs, antibiotics and surgical resection can reduce symptoms (1).

The etiology of HS is poorly understood (3). While mutations in γ-secretase genes were detected in a small subset of HS patients, their link with the pathophysiology of HS has remained elusive (4). For the large majority of cases, HS is currently believed to involve defects in genes regulating inflammation and immunity (5). In support of this view, persistent production of pro-inflammatory Interleukin (IL)-1β is a distinctive feature of HS lesions (6–8). Tumor Necrosis Factor (TNF)-α is also frequently induced in advanced HS lesions and the TNF-α blocker adalimumab had a positive effect on the clinical outcome of moderate-to-severe HS (9). Infiltrating myeloid cells are typically found in skin lesions, with antimicrobial peptide and interferon (IFN) signatures reflecting the generation of potent innate immune responses to infection (6, 10–15). An aberrant activation of neutrophils and B cells was also reported, which may jointly contribute to perpetuate lesion inflammation in patients with HS (12, 13). Finally, T cells recruited to HS skin lesions produce IL-17A and IFN-γ but little IL-22 (6, 7, 10, 16-19), an immune signature contrasting with the increased frequency of Th17 and Th22 cells, but not Th1 cells in peripheral blood (6, 10-13, 19).

In parallel with dysregulated immune responses, metagenomic analyses have identified a distinctive anaerobic microbiota in HS skin, the expansion of which correlates with lesion severity (20, 21). Antibiotic combinations targeting these anaerobic bacteria efficiently eliminated HS symptoms, and remission could be prolonged with monotherapy maintenance treatments (22–26). Interestingly, inflammatory bowel disease (IBD) and metabolic syndrome are frequent HS co-morbidities (27–30). In both of these conditions, unresolved inflammation was linked to perturbations of the gut microbiota-host metabolic homeostasis. We therefore hypothesized that metabolic alterations may link the cutaneous dysbiosis and immune dysregulation characterizing HS pathophysiology. This prompted us to conduct the first metabolomic analysis of HS skin.

## Results

### The kynurenine pathway of tryptophan catabolism is activated in HS skin lesions

To capture the biochemical changes coinciding with the most frequent form of the disease, we compared the metabolomes of paired lesional and clinically normal skin samples from patients with Hurley Stage I HS (Table S1). Biopsy specimens were also harvested from healthy subjects, in order to detect constitutive alterations in HS skin metabolism. Our semi-quantitative analysis detected a total of 500 metabolites, which distributed differently among skin samples from healthy controls (HC), healthy and lesional skin from HS patients (H-HS and L-HS, respectively) (Fig. 1A). A principal component analysis clearly discriminated the metabolomes of HC and L-HS, while H-HS samples showed an intermediate profile (Fig. 1B).

**Figure 1.**
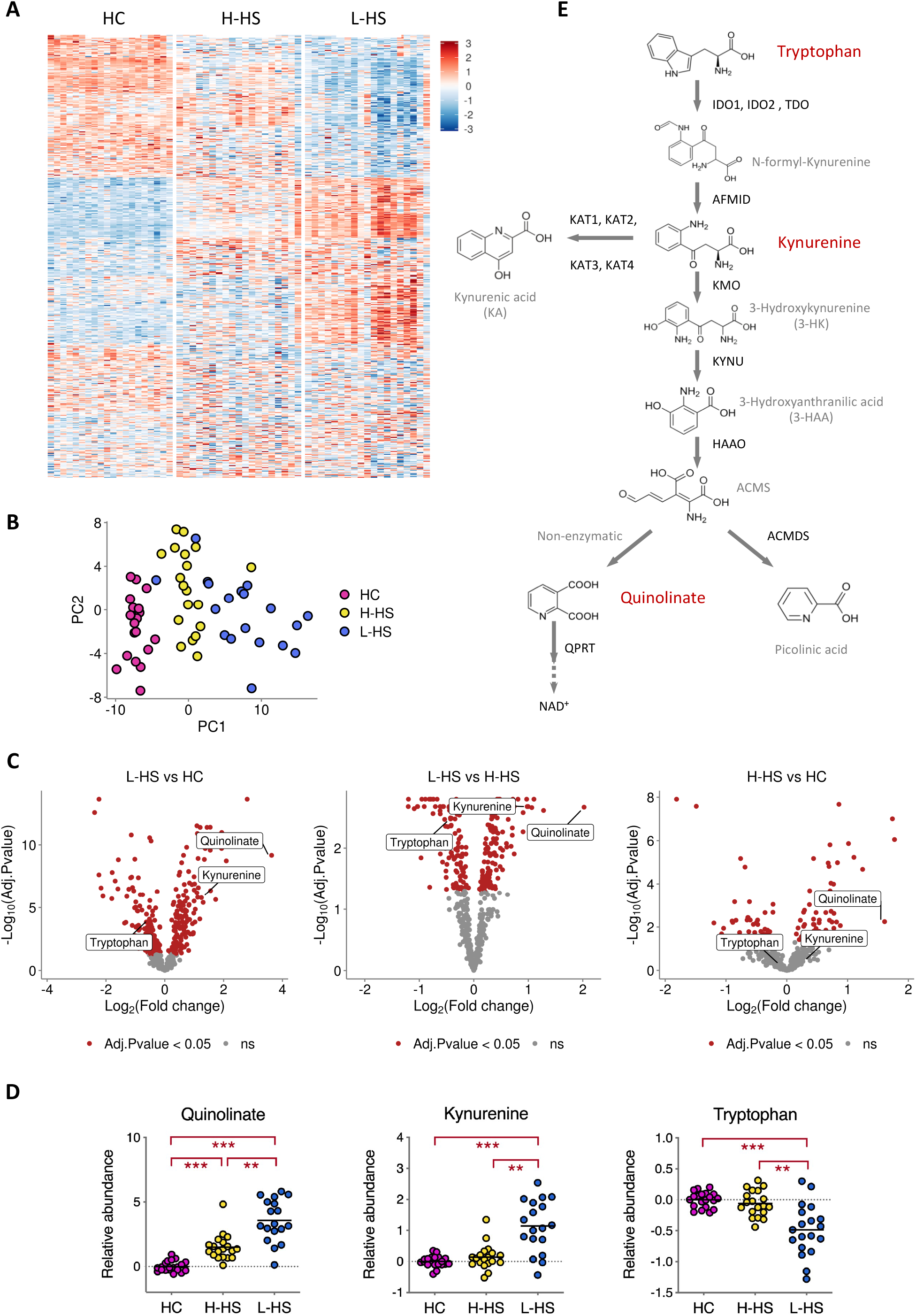
The kynurenine pathway of Trp catabolism is activated in HS skin lesions. (**A**) Heatmap showing the relative abundance of metabolites in skin biopsies from HC and HS patients (H-HS, healthy skin; L-HS, lesional skin). (**B**) Principal component analysis (PCA) based on metabolite abundance in skin samples. Axes correspond to the 2 first principal components. (**C**) Volcano plots illustrating pairwise comparisons of relative metabolite levels between groups. (**D**) Relative levels of Trp metabolites in skin samples from HC and HS patients, displayed as scatter dot plots with means. \**P*<0.05, \*\**P*<0.01, \*\*\**P*<0.001 by Welch’s two-sample t-test for comparisons with HC and matched pairs t-test for comparisons of L-HS with matched H-HS, with multiple testing correction. (**E**) Kynurenine pathway of Trp catabolism.

We found that 335 metabolites were differently modulated between L-HS and HC, among which 165 were significantly augmented in L-HS (Table S2). Notably the most up-regulated metabolite in L-HS, compared to both HC and H-HS, was quinolinate (Quin) (Fig. 1C and 1D). Quin levels were also significantly up-regulated in H-HS, compared to HC (Fig. 1C and 1D). Quin is a downstream product of the kynurenine (Kyn) pathway of tryptophan (Trp) catabolism culminating in nicotinamide adenine dinucleotide (NAD+) production (Fig. 1E). Like Quin, Kyn was significantly up-regulated in L-HS compared to both H-HS and HC (Fig. 1C and 1D). Conversely, Trp levels were decreased in L-HS, compared to both H-HS and HC (Fig. 1C and 1D). Altogether, these data revealed constitutive alterations in HS skin metabolism, with accumulation of Kyn and Quin in lesions suggesting a local activation of Trp catabolism via the kynurenine pathway.

### Distinctive and selective induction of kynurenine pathway enzymes in HS skin

To determine whether the observed alterations in Trp metabolism were associated with changes in expression of kynurenine pathway enzymes, a new cohort of Hurley Stage I HS patients and healthy controls was recruited for transcriptomic analysis of skin biopsies (Table S1). The first step of Trp degradation by the kynurenine pathway is regulated by 3 rate-limiting enzymes: Indoleamine 2,3-dioxygenases (IDO)1, IDO2 and Trp 2,3-dioxygenase (TDO) (Fig. 2A). While *IDO2* was poorly expressed in all skin samples (data not shown), *IDO1* and *TDO* transcripts were both detected and the two genes were more highly expressed in L-HS than in HC (Fig. 2B). Downstream of IDO1 and TDO, Kyn can be converted into 3-Hydroxykynurenine (3-HK) by KMO or Kynurenic acid (KA) by KAT enzymes (Fig. 2A). Expression of *KMO* was augmented in L-HS while that of *KAT1* was decreased (Fig. 2B), suggesting that Kyn is preferentially metabolized into Quin in HS skin lesions. Downstream of KMO, expression of *KYNU* (converting 3-HK into Hydroxyanthranilic acid, 3-HAA) was also augmented in L-HS compared to HC (Fig. 2A-B). In contrast, that of *HAAO* (immediately downstream of KYNU) and *QPRT* (converting Quin into a NAD precursor) were not significantly modulated (Fig. 2A-B). The potent induction of *IDO1, TDO, KMO and KYNU* without concomitant upregulation of *KAT1* and *QPRT* was thus consistent with the decreased levels of Trp and the accumulation of Kyn and Quin in HS skin lesions (Fig. 1D).

**Figure 2.**
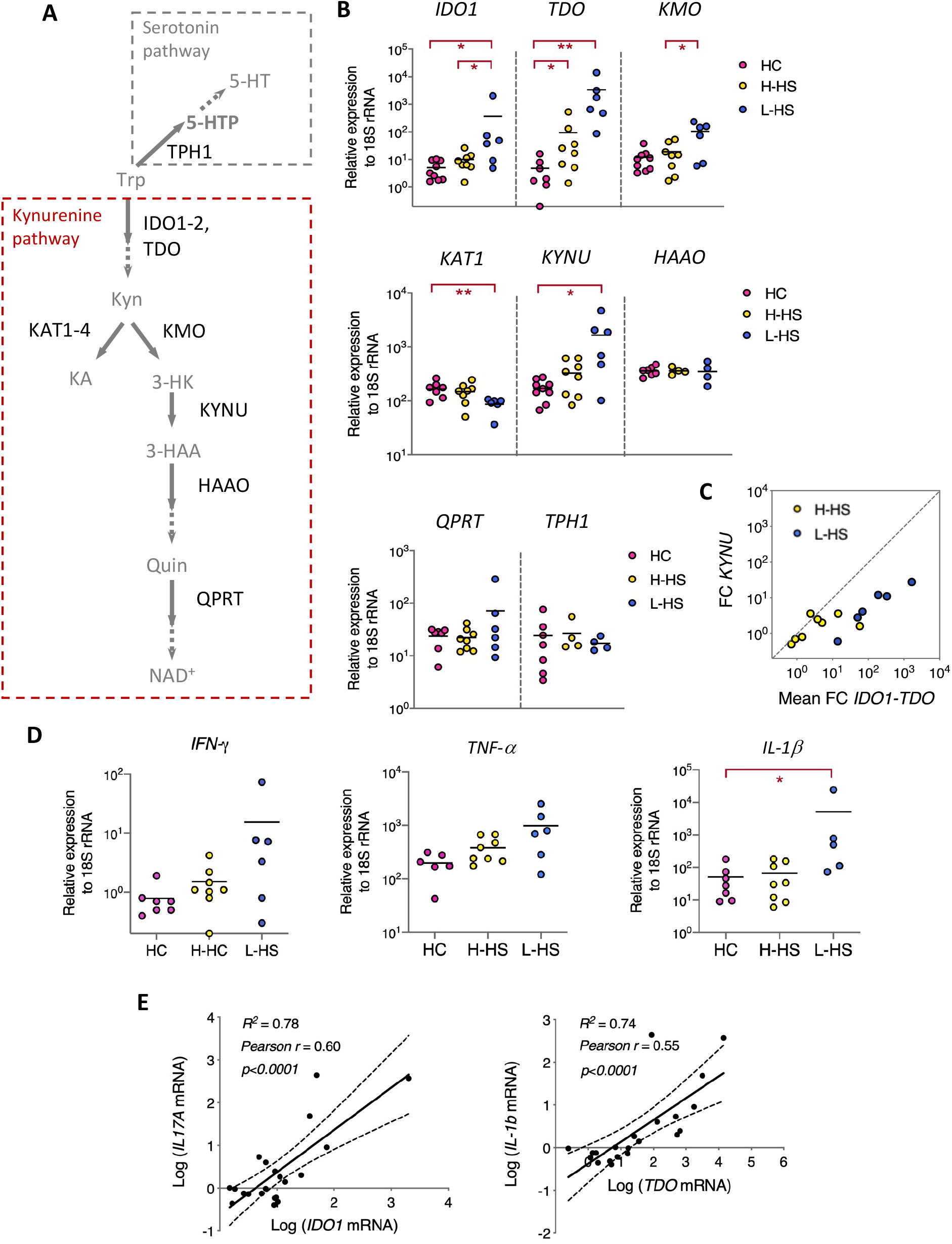
Distinctive and selective induction of kynurenine pathway enzymes in HS skin. **(A)** Enzymatic steps in the kynurenine and serotonin pathways of Trp catabolism. **(B)** Relative expression of kynurenine pathway enzymes in skin samples from HC and HS patients. Data are shown as scatter dot plots with means. \**P*<0.05, *P*\*\*<0.01, \*\*\**P*<0.001 by Mann-Whitney U-test for comparisons with HC, and Wilcoxon matched-pairs test for comparisons of L-HS with matched H-HS. **(C)** Fold changes (FC) in expression of *IDO* and *TDO* (mean FC of the two genes) and *KYNU* are shown for H-HS (yellow) and L-HS (blue) skin samples. (**D**) Relative expression of cytokines in skin samples from HC and HS patients, shown as scatter dot plots with means. \**P*<0.05, \*\**P*<0.01 by Mann-Whitney U-test for comparisons with HC, and Wilcoxon matched-pairs test for comparisons of L-HS with matched H-HS. (**E**) Correlation between *IDOl/IFN-γ* and *TDO/IL-1β* gene expression. Linear regression plots are shown, with R squared (R2), Pearson coefficient (r) and statistical significance.

Previous work has identified *KYNU* as one of the most upregulated genes in lesional skin of patients with psoriasis and atopic dermatitis (31–34), and preferential upregulation of *KYNU* over *IDO1* and *TDO* corelated with disease severity in these settings (35). In HS skin, *IDO1* and *TDO* were more highly expressed than *KYNU*, particularly in lesions (Fig. 2C), indicating a distinct activation pattern of the KP in this context that is not generically related to skin inflammation.

In addition to its catabolism to NAD, Trp can serve as a precursor for the neurotransmitter serotonin (5-HT), via conversion into 5-HTP by Trp hydroxylase 1 enzyme (TPH1) (Fig. 2A) (36). Our metabolomic analysis did not detect any product of the serotonin pathway and *TPH1* was comparably expressed in skin from HS patients and controls (Fig. 2B), suggesting that this pathway was not modulated in HS skin. Together, our data in Figures 1 and 2A-B thus revealed a distinctive and selective induction of the kynurenine pathway of Trp catabolism in HS skin lesions.

IDO1 and TDO are important links between the kynurenine pathway and inflammation, because their production is initiated by pro-inflammatory cytokines. IFN-γ is a powerful driver of *IDO1* gene expression and IFN-γ-mediated induction of *IDO1* can be potentiated by other cytokines such as TNF-α or IL-1β, while IL-1β itself is a *TDO* inducer (37, 38). Although statistical significance was not reached, expression of both *IFN-γ* and *TNF-α* tended to increase in lesions and *IL-1β* expression was significantly augmented in L-HS, compared to HC (Fig. 2D). Notably, there was a strong positive correlation between *IDO1* and *IFN-γ* on the one hand, and *TDO* and *IL-1β* on the other hand (Fig. 2E), suggesting that IFN-γ and IL-1β produced by infiltrating immune cells locally activate Trp catabolism *via* transcriptional induction of *IDO1* and *TDO*.

### Immune infiltrates and dermal fibroblasts both contribute to Quin production in HS skin

We next used multi-color immunohistochemistry to determine which cells were responsible for Quin production in HS skin. No Quin staining was detected in the epidermis of HS patients or controls (data not shown). However, in line with our metabolomic data, Quin+ cells were found in the dermis of HS skin, and lesions contained significantly higher number of Quin+ cells compared to non-lesional and normal skin. Notably, the frequency of Quin+ cells in H-HS was superior to HC (Fig. 3A), further indicating that Quin production is altered in HS skin in the absence of lesion.

**Figure 3.**
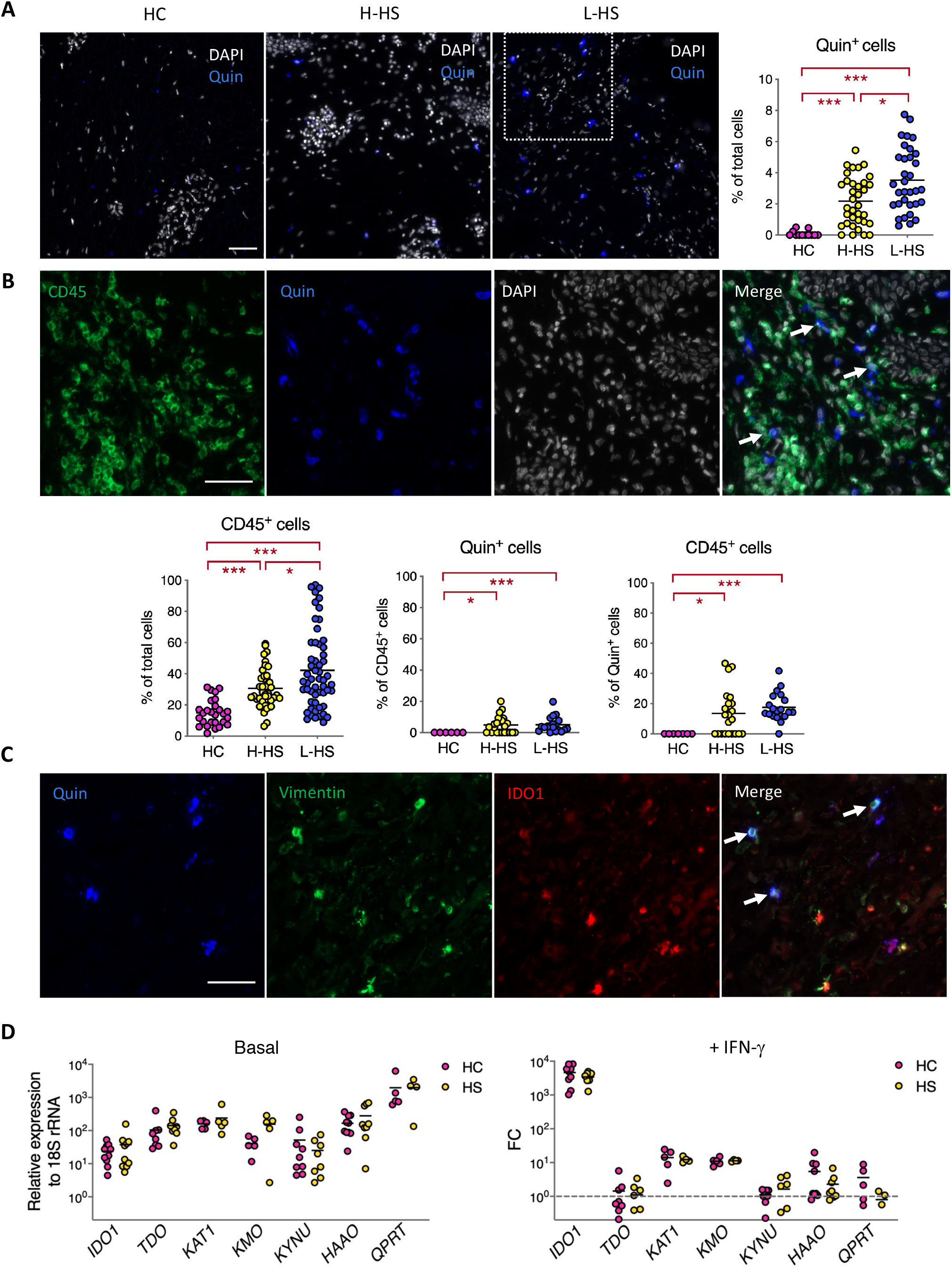
Immune infiltrates and dermal fibroblasts jointly contribute to Quin production in HS skin. (**A**) DAPI and Quin staining of representative HC, H-HS and L-HS dermis sections, with mean incidence of Quin+ cells, relative to total DAPI+ cells, as measured on >5 skin sections from 3 HC and 4 HS patients. (**B**) CD45, Quin and DAPI staining of a representative L-HS dermis section, with arrows pointing to Quin+ CD45+ immune cells; Quantification of CD45+ cells relative to total DAPI+ cells, CD45+ cells amongst Quin+ cells and Quin+ cells amongst CD45+ cells. (**C**) Zoomed view of the area depicted in (A), following Quin, Vimentin and IDO1 staining, with arrows pointing to Quin+ Vimentin+ IDO1+ cells. (**D**) Relative levels of kynurenine pathway enzyme transcripts in primary fibroblasts from 5 HC and 5 HS patients in resting conditions (left); Fold change (FC) in gene expression following a 24h treatment with IFN-γ (2.5 ng/ml), relative to unstimulated controls (right). Scale bar 50 μm (A, B, C). Scatter dot plots with means, * *P* <0.05, \*\**P*<0.01, \*\*\**P*<0.001 by Mann-Whitney U-test, with multiple testing correction.

Since cytokine-induced expression of IDO can occur in immune cells (37), the production of Quin by these cells was first investigated. Lesional skin, and non-lesional skin to a lesser extent, displayed significant infiltration of CD45+ hematopoietic cells and a fraction of these cells stained positive for Quin (Fig. 3B). However, CD45+ hematopoietic cells only represented <20% Quin+ cells in L-HS (Fig. 3B), and >50% of Quin+ cells in L-HS were found to express the fibroblast marker Vimentin (Fig. 3C). Dermal fibroblasts are known to degrade Trp in response to stimulation with IFN-γ via induction of IDO1 (39). Consistent with this finding, Quin+ Vimentin+ cells in L-HS stained positive for IDO1 (Fig. 3C). To determine if fibroblasts of HS patients were intrinsically prone to degrading Trp, we generated primary fibroblasts from patients and controls (Table S1) and analyzed their expression of kynurenine pathway enzymes. There was no significant difference in basal expression of *IDO1*, *TDO, KAT1, KMO*, *KYNU*, *HAAO* and *QPRT* in fibroblasts of HS patients or controls, and HS fibroblasts responded normally to IFN-γ, TNF-α and IL-1β stimulation (Fig. 3D and Fig. S1). We concluded that enhanced production of Quin in L-HS does not result from a constitutive activation of the kynurenine pathway, but rather from cytokine-driven induction of Trp catabolism in dermal fibroblasts and a subset of infiltrating immune cells.

### Plasma Trp levels are decreased in HS patients

The data in Figures 1-3 revealed alterations in the Trp metabolism of HS skin that are associated with the local production of inflammatory cytokines. To determine if these immune and metabolic perturbations extended beyond skin lesions, we examined the production of kynurenine pathway-inducing cytokines by peripheral blood lymphocytes (PBLs). Consistent with previous studies, PBLs from HS patients showed an increased frequency of IL-17A and IL-22-producing T cells, while IFN-γ-producing T cells were unaltered (Fig. 4A and Fig. S2A). While IL-17A protein levels were significantly augmented in the plasma of HS patients, IFN-γ levels remained unchanged (Fig. 4B). Moreover, TNF-α and IL-1β plasma levels were below the detection limit in all samples (data not shown), thus excluding a systemic inflammatory syndrome in the patients studied.

**Figure 4.**
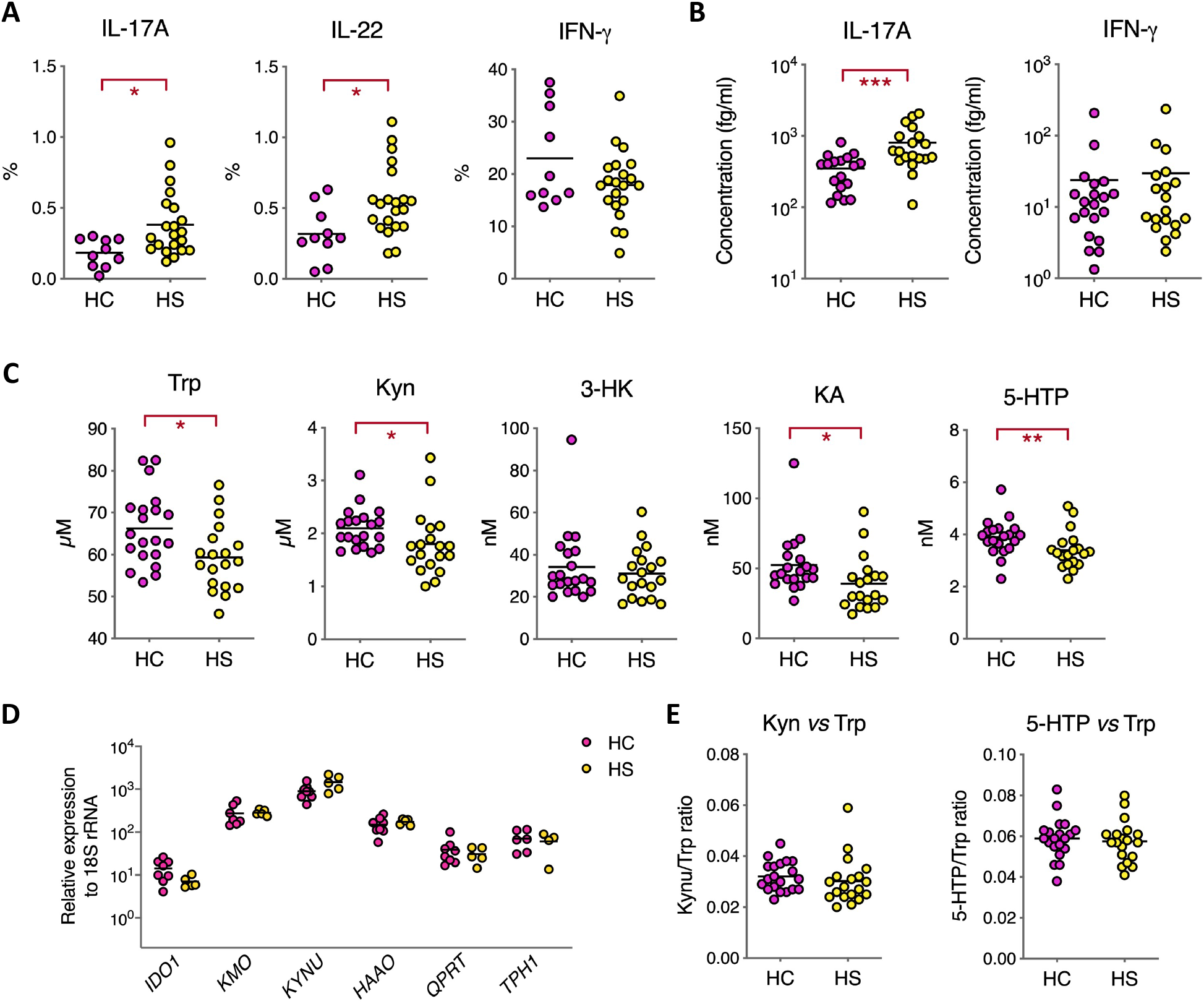
Plasma Trp levels are decreased in HS patients. (**A**) Frequency of cytokine producing blood cells, relative to total T cells, following a 3h stimulation with PMA/Ionomycin. (**B**) Cytokine concentrations in plasmas from HC and HS patients. (**C**) Comparison of Trp metabolite concentrations in plasma of HC and HS patients. (**D**) Relative expression of kynurenine pathway enzymes in PBMCs from HC and HS patients. (**E**) Ratios of Kyn or 5-HTP to Trp in plasma. Data are shown as scatter dot plots with means. \**P*<0.05, \*\**P*<0.01, \*\*\**P*<0.001 by Mann-Whitney U-test.

We next investigated if Trp metabolism was altered at the systemic level by quantifying Trp metabolites in the plasma. Plasma levels of Trp, Kyn, KA and 5-HTP were significantly lower in HS patients, compared to controls (Fig. 4C). The decrease in plasma levels of Trp metabolites was not due to differential induction of the kynurenine or serotonin pathways in peripheral blood mononuclear cells (PBMCs), as the basal expression of *IDO1*, *KMO*, *KYNU*, *HAAO*, *QPRT* and *TPH1* was comparable in PBMCs from HS patients and controls (Fig. 4D). Moreover, the Kyn/Trp and 5-HTP/Trp ratios were unchanged (Fig. 4E). In conclusion, this analysis revealed abnormally low levels of circulating Trp in HS patients that were not accompanied by a systemic inflammatory state.

### Activation of AHR is defective in HS skin lesions

Our observation that Trp levels were decreased in HS skin lesions (Fig. 1D) suggested that Trp catabolism by resident bacteria may be altered. This essential amino acid is indeed an energy source for the microbiota, which modulates local immune responses through production of indole and derivatives (40). Microbiota-derived indole metabolites include ligands of the Aryl Hydrocarbon Receptor (AHR), a transcription factor translating metabolic signals into cell typespecific gene expression programs at mucosal surfaces (41) (Fig. 5A). All cells in the skin express AHR and physiological activation of AHR in keratinocytes and dermal fibroblasts is critical for the regulation of skin inflammatory responses (40). To determine whether AHR signaling was altered in HS skin, we compared the expression of *AHR* and *AHR*-dependent genes in healthy and diseased skin samples. While *AHR* expression was comparable in HC, H-HS and L-HS, that of AHR target genes *AHRR*, *CYP1A1* and *CYP1A2* was significantly decreased in L-HS, compared to HC (Fig. 5B). To see if defective activation of AHR in HS skin lesions was due to intrinsic defects in AHR signaling, we next examined the integrity of this pathway in fibroblasts of HS patients. Figure 5C shows that HS patients- and HC-derived fibroblasts displayed comparable expression of *AHR, AHRR and CYP1B1* in the resting state; and when stimulated with the prototypical AHR agonist FICZ, displayed comparable induction of *AHRR* and *CYP1B1*. This suggested that decreased activation of AHR in lesional skin is not due to intrinsic defects, but rather caused by locally impaired production of AHR agonists. Known agonists of AHR include Kyn, a low-affinity ligand whose physiological relevance is doubtful (42), and indole derivatives originating from bacterial degradation of Trp. Amongst bacterial metabolites of Trp, our metabolomic analysis detected indole-3-acetic acid (IAA), indole-3-lactic acid (ILA) and indoxylsulfate (IS), of which IAA is a recognized AHR agonist (43). Notably, IAA levels were significantly decreased in L-HS, compared to H-HS and HC (Fig. 5D). Therefore, the defective activation of AHR in HS skin lesions coincides with an impaired production of AHR agonist IAA by the skin microbiota.

**Figure 5.**
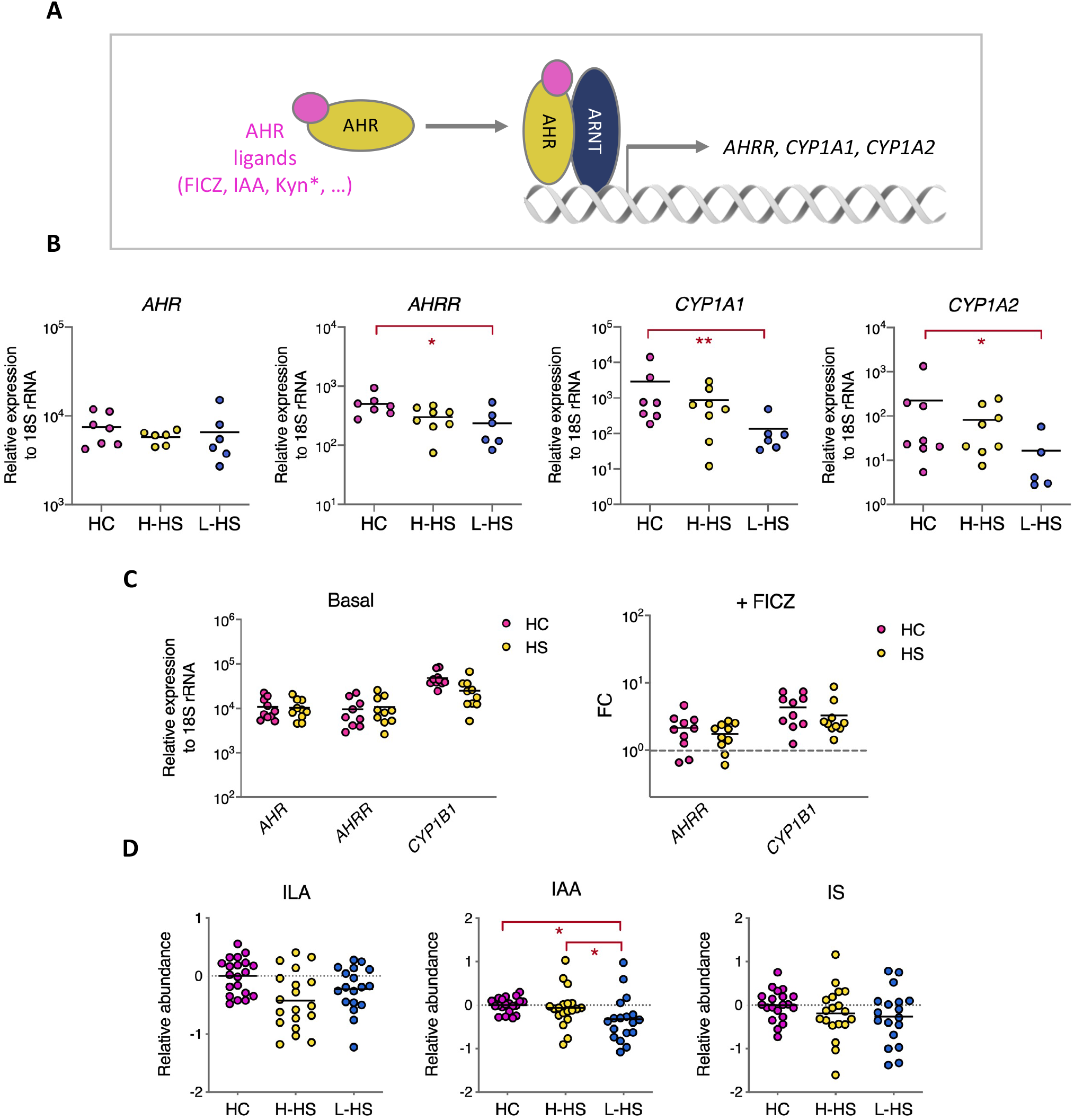
Activation of AHR is defective in HS skin lesions. (**A**) The AHR pathway. Upon ligand binding, AHR translocates into the nucleus where association with ARNT and interaction with specific genomic sequences induce the transcription of target genes (*active in supra-physiological concentrations). (**B**) Relative levels of AHR and AHR-controlled transcripts in skin samples from HC and HS patients. \**P*<0.05, \*\**P*<0.01 by Mann-Whitney U-test for comparisons with HC, and Wilcoxon matched-pairs test for comparisons of L-HS with matched H-HS. (**C**) Relative expression of AHR-controlled transcripts in fibroblasts from HC and HS patients in resting (left) or FICZ-stimulated (right) conditions. Data are from 2 independent experiments with fibroblasts from 5 HC and 5 HS patients. (**D**) Relative levels of indole metabolites in skin samples from HC and HS patients. \**P*<0.05, \*\**P*<0.01, by Welch’s two-sample t-test for comparisons with HC and matched pairs t-test for comparisons of L-HS with matched H-HS, with multiple testing correction.

## Discussion

In this report, we reveal that patients with Hurley Stage I HS display local and systemic alterations in Trp metabolism. A selective and potent induction of the kynurenine pathway of Trp degradation was detected in HS skin lesions. Based on our histological (Fig. 3C,D) and transcriptomic (Fig. 2B,D and S1) data, we propose that induction of this pathway results from the recruitment and activation of immune cells, whose release of IFN-γ and IL-1β jointly contribute to trigger expression of *IDO1* and *TDO* in dermal fibroblasts and a subset of infiltrating immune cells. The alterations in Trp metabolism that we observed in HS skin differ from those previously reported in other inflammatory skin diseases, in magnitude and profile. In HS skin lesions transcriptional activation of the kynurenine pathway was dominated by expression of *IDO1* and *TDO*, leading to local accumulation of Kyn and Quin (Fig. 1-2). These two enzymes were either not modulated or only mildly up-regulated in published transcriptomes of atopic and psoriatic skin (31–34), and the skin levels of kynurenine pathway metabolites were unchanged in these settings (44, 45). While *KYNU* is one of the top up-regulated genes in both atopic dermatitis and psoriasis, induction of *KYNU* was lower than that of *IDO1/TDO* in HS skin (Fig. 2C). Differently from *IDO1*, *KYNU* expression is optimally triggered by the combination of IFN-γ and TNF-α (35). Its relatively lower induction in HS skin lesions may reflect a comparatively less important production of TNF-α in studied patients. Since TNF-α potentiates both the IFN-γ-driven-induction of *IDO1* and expression of *KYNU*, its upregulation in more severe HS lesions may further activate the kynurenine pathway. It is important to note that our study is the first to focus on a clinically homogenous group of Hurley Stage I patients. This might explain differences with previous studies, which involved patients with variable and often severe HS lesions (10, 12, 14, 15). Our immune profiling of PBMCs from HS patients highlighted higher frequencies of NK cells expressing Granzyme B and Perforin and producing IFN-γ upon stimulation (Fig. S2B-D). Since NK cells are potent producers of IFN-γ that were identified in HS lesions (Fig. S2E), these data suggest that NK cells are primed for effector functions in HS patients, and that their local activation may contribute to *IDO1* induction in lesional skin.

Importantly, our analysis of HS plasmas also revealed for the first-time abnormally low levels of circulating Trp (Fig. 4C). Whether depletion in plasma Trp results from increased degradation in HS skin or from metabolic dysregulations in other organs is an open question. Since Trp is an essential amino acid, our data suggest that intestinal absorption or liver catabolism of Trp may be altered in HS patients.

We found that CD45+ Quin+ cell numbers, Quin levels and *TDO* transcripts were elevated in clinically normal skin of HS patients (Fig. 1D, 2B and 3B). This suggests that inflammation and kynurenine pathway are induced throughout HS skin, with the caveat that H-HS skin samples were harvested in the periphery of lesions in the present work. Recent metagenomic studies have revealed a distinctive dysbiosis in clinically normal skin of HS patients, marked by the expansion of *Corynebacterium* spp and anaerobes (46). In line with these findings, our analysis of the skin microbiota using 16S ribosomal RNA sequencing showed that the microbiome of H-HS skin differed significantly from that of HC by the increased abundance of Corynebacterium spp and anaerobic bacteria (Fig. S3A-B). Expansion of these bacterial species may contribute to stimulate inflammation and Trp degradation at a pre-clinical stage of disease.

Over-activation of the kynurenine pathway could contribute to the underlying pathogenic mechanisms of HS in various ways. First, decreased availability of Trp in the skin may provide Trp-independent pathobiont such as *Staphylococcus aureus* with a selective growth advantage. A recent analysis of skin microbial communities coupled to cutaneous gene expression in patients with atopic dermatitis or psoriasis found that dense colonization by *S. aureus* correlates with increased expression of *TDO, KYNU* and *KMO* (31). While our analysis of HS skin microbiota failed to identify *Staphylococci* to species level, *S. aureus* was previously identified as an important colonizer of HS skin lesions with a prevalence rate reaching up to 56% (47). Our findings suggest that activation of the kynurenine pathway in HS skin lesions may favor the expansion of this pathobiont.

Second, decreased Trp availability may lower the production of AHR ligands by the skin microbiota, thereby altering AHR-mediated regulation of inflammation. Amongst bacterial AHR agonists, our metabolomic analysis detected the *Lactobacillus* product IAA. Our observation that IAA was decreased in lesional skin, while Kyn and Quin were instead increased, indicates that Trp degradation by *Lactobacilli* was downmodulated. In support of this hypothesis, the relative abundance of *Lactobacillus* spp was significantly lower in L-HS compared to HC (Fig. S3C). Recent studies of atopic skin have reported a selective decrease in skin levels of the IAA-product indole-3-aldehyde (IAId) (45). Notably, treating with IAId topically or orally alleviated skin inflammation in a mouse model of atopic dermatitis, in a AHR dependent manner (45). Altogether, these data strongly support the view that defective production of AHR agonists by resident bacteria contributes to immune dysregulation in HS skin lesions.

Finally, depression and anxiety are common conditions in patients with HS (2). Over-activation of the IDO1 pathway in patients with chronic inflammatory diseases, or in patients treated with type I IFN was proposed to cause depression via central serotonin depletion or increased production of neurotoxic Quin in the brain (36). Plasma levels of Trp, Kyn and 5-HTP were significantly reduced in HS patients (Fig. 4C). Since these Trp metabolites are able to cross the blood-brain-barrier, dysregulated Trp metabolism in HS skin could impact brain chemistry and predispose to depressive symptoms.

In conclusion, this study reports for the first time local and systemic alterations in the Trp metabolism of HS patients. Dysregulation of Trp catabolism at the host-skin microbiota interface provides a potential mechanism for the diverse clinical manifestations of HS. In the absence of animal models for HS, further investigations in patients will be needed to elucidate the molecular origin of these metabolic defects. Trp degradation via the kynurenine pathway has immunosuppressive effects, operating through the formation of catabolites inducing a regulatory phenotype in T cells and DCs (41). However, there is accumulating evidence that in barrier organs such as the gut and the skin, exacerbated IDO activity may downregulate AHR agonist production by the microbiota, thereby promoting chronic inflammation (36, 40). While Trp metabolites generated by the gut microbiota are now well characterized (48), little is known about Trp metabolism by skin-resident bacteria and their capacity to generate immunomodulatory AHR ligands. Correcting AHR signaling via administration of AHR agonist FICZ or transplantation of Trp-metabolizing *Lactobacillus* strains proved to be effective at reducing intestinal inflammation in a mouse model of IBD (49), and FICZ also attenuated skin inflammation in a mouse model of psoriasis (50). Studies on the effects of topically-applied bacterial products and skin bacterial transplants have yielded promising results in animal models and human studies of atopic dermatitis (45, 51). Our study suggests that such approaches may help restore immune homeostasis in HS skin.

## Methods

### Study design and harvested specimens

The characteristics of HS patients and healthy individuals recruited for this study are listed in Table S1. Specific inclusion criteria were age ≥ 18 years and, for patients, diagnosis of active Hurley stage 1 HS. Non-inclusion criteria for both patients and healthy subjects were: pregnancy, antibiotic or immunosuppressive treatment during the past month, chronic inflammatory disease, cancer, hematological malignancy and contraindication to biopsy. For healthy subjects, a progressive skin disease, or any personal or familial history of chronic inflammatory disease constituted additional non-inclusion criteria.

### Harvested specimens

In patients with HS, punch biopsies (4 mm diameter) were harvested in lesional skin and clinically normal, peri-lesional skin (approximately 5 cm from the inflamed nodule). Biopsy specimens from HC were harvested in the armpit area. Immediately after harvesting, biopsies were snap frozen at −80°C until metabolomic, transcriptomic (in RNAlater) and histologic analysis. Primary fibroblasts were derived from healthy peri-lesional skin (HS patients) or normal skin from surgical margins (controls) as described previously (52). Primary fibroblasts were cultivated in DMEM (Gibco) supplemented with 10% fetal calf serum and 1% Penicillin-Streptomycin and stimulated for 24 h with 100 ng/ml of the AHR agonist FICZ, or with IFN-γ (2.5 ng/ml) in the presence or absence of TNF-α (100 ng/ml). Venous blood (2 × 10 mL) was collected and within 1 h, PBMCs and plasmas were separated by Ficoll and frozen at −196°C/−80°C, respectively until proteomic flow cytometric studies.

### Metabolomic profiling of skin biopsies

Skin samples were prepared using the automated MicroLab STAR® system (Hamilton Company) for global untargeted metabolic profiling by Metabolon Inc. (Durham, NC). Proteins were precipitated with methanol under vigorous shaking for 2 min (Glen Mills GenoGrinder 2000) followed by centrifugation. The resulting extract was divided into five fractions: two for analysis by two separate reverse phase (RP)/UPLC-MS/MS methods with positive ion mode electrospray ionization (ESI), one for analysis by RP/UPLC-MS/MS with negative ion mode ESI, one for analysis by HILIC/UPLC-MS/MS with negative ion mode ESI and one sample was reserved for backup. Samples were placed briefly on a TurboVap® (Zymark) to remove the organic solvent. Ultra-performance liquid chromatography (UPLC)- mass spectrometer (MS)/MS analysis of the resulting samples used a Waters ACQUITY UPLC and a Thermo Scientific Q-Exactive high resolution/accurate MS interfaced with a heated electrospray ionization (HESI-II) source and Orbitrap mass analyzer operated at 35,000 mass resolution. The sample extract was dried then reconstituted in solvents compatible to each of the four methods. Each reconstitution solvent contained a series of standards at fixed concentrations to ensure injection and chromatographic consistency. One aliquot was analyzed using acidic positive ion conditions, chromatographically optimized for more hydrophilic compounds. In this method, the extract was gradient eluted from a C18 column (Waters UPLC BEH C18-2.1×100 mm, 1.7 μm) using water and methanol, containing 0.05% perfluoropentanoic acid (PFPA) and 0.1% formic acid (FA). Another aliquot was also analyzed using acidic positive ion conditions, however it was chromatographically optimized for more hydrophobic compounds. In this method, the extract was gradient eluted from the same afore mentioned C18 column using methanol, acetonitrile, water, 0.05% PFPA and 0.01% FA and was operated at an overall higher organic content. Another aliquot was analyzed using basic negative ion optimized conditions using a separate dedicated C18 column. The basic extracts were gradient eluted from the column using methanol and water with 6.5 mM Ammonium Bicarbonate, pH 8. The fourth aliquot was analyzed via negative ionization following elution from a HILIC column (Waters UPLC BEH Amide 2.1×150 mm, 1.7 μm) using a gradient consisting of water and acetonitrile with 10mM Ammonium Formate, pH 10.8. The MS analysis alternated between MS and data-dependent MSn scans using dynamic exclusion. The scan range varied slightly between methods but covered 70-1000 m/z. Subsequent bioinformatic analysis used Metabolon proprietary tools for raw data extraction, peak-identification software, QC and compound identification. A total of 500 biochemicals were identified in our dataset. Metabolite data were log2-transformed and normalized using Pareto scaling. The statistical analysis was performed on R software using the limma package (53). False discovery rates (FDR) were calculated using the Benjamini-Hochberg method (Table S2).

### Quantitative assay of Trp metabolites in serum

Blood levels of five metabolites (Trp, Kyn, 3-HK, KA and 5-HTP) were assessed in serum samples by high-performance liquid chromatography coupled to tandem mass spectrometry, using methodologies adapted from (54).

### PBMCs studies

Ficoll-isolated PBMCs were activated with PMA (10 ng/ml; Sigma-Aldrich) plus ionomycin (1 μg/ml; Sigma-Aldrich) during 3 h at 37°C in the presence of BD GolgiPlug (brefeldin A; BD Biosciences) and BD GolgiStop (monensin; BD Biosciences). Flow cytometry analysis of T cells and NK cells was performed using the following antibodies. Biotinylated anti-human CD1a (HI149), CD3 (OKT3), CD14 (61D3), CD19 (HIB19), CD34 (4H11), CD123 (6H6), CD203c (FR3-16A11), CD303 (AC144), TCRαβ (IP26), TCRγδ (B1) and FcεRIα (AER-37) Abs in combination with the streptavidin BV711; and conjugated anti-human CD5 Alexa Fluor 700 (L17F12), CD7 BV650 (M-T701), CD56 BUV737 (NCAM16.2), CD127 PE-Cy7 (eBioRDR5), NKp44 BB515 (p44-8), CD69 PE (FN50), PD-1 BV786 (EH12.2H7), CD25 BUV563 (2A3), HLA-DR BUV661 (G46-6), CD45 BUV805 (HI30), EOMES PE-eFluor 610 (WD1928), Perforin BV421 (dG9), Granzyme B PE-CF594 (GB11), IFN-γ BUV395 (B27), IL-17A BV570 (BL168) and IL-22 PE (22URTI) antibodies were purchased from Biolegend, ThermoFisher, BD Biosciences or Miltenyi Biotec. Fc receptors were blocked using IgG from human serum (Sigma-Aldrich). Surface membrane staining was performed in Brilliant Stain Buffer (BD Biosciences). Transcription factors and cytokines were stained using the Foxp3 staining buffer set (ThermoFisher) according to the manufacturer’s instructions. The fixable viability dye eFluor 506 (ThermoFisher) was used to exclude dead cells. Samples were acquired on a Symphony A5 (BD Biosciences) using FACSDiva 8 and analyzed with FlowJo 10 (BD Biosciences).

### Quantitative reverse-transcription PCR (qRT-PCR)

Total RNA was extracted from pulverized skin biopsies or cell pellets with Qiazol lysis reagent (Qiagen), then purified using Qiagen RNeasy Mini Kit and digested with RNase-Free DNase set (Qiagen 79254) for 15 min at room temperature. First-strand cDNA was synthesized from 400ng of total RNA with the high capacity cDNA reverse transcription kit (Applied Biosystems 4368814). Expression was quantified using Power SYBR Green PCR Master Mix (Applied Biosystems 4367659) and gene-specific primers (Table S3). Amplification was performed from 5 ng of cDNA template in a final volume of 20 μl. in a 96-well PCR plate. Amplification conditions were 2 min at 50 °C, 10 min at 95 °C, followed by 40 cycles of 15 s at 95 °C and 1 min at 60 °C on a Quant Studio 3 Real-time PCR System (Applied Biosystems). Results were normalized by relative expression using 18S rRNA as an endogenous control.

### Histology

Frozen skin biopsies were fixed overnight at 4°C with 4% paraformaldehyde in phosphate buffer. Tissues were cryopreserved by immersion in 30% sucrose for 24 h then embedded in optimal cutting temperature compound (Tissue-Tek O.C.T, Sakura) and stored at −80°c until sectioning. Immunostaining was performed on 8 μM sections obtained with a Cryostat (CM3050 S, Leica) onto SuperFrost Plus adhesion slides (ThermoFisher). Non-specific staining was blocked 1 h with 0.1% Triton, 10% fetal-calf serum before overnight incubation with primary antibodies at 4°C. Primary antibodies were rabbit polyclonal anti-Quinolinic acid (Abcam, ab37106), mouse monoclonal anti-human CD45 (2D1) (BD Biosciences, 347460), Rabbit polyclonal anti-human CD3s (Dako, A0452), sheep polyclonal anti-human CD7 (R&D Systems, AF7579), mouse anti-IDO (1F8.2) (Merck, MAB10009) and polyclonal goat anti-Vimentin (R&D Systems, AF2105). Donkey anti-rabbit A647 (ThermoFischer Scientific, A-31573), anti-sheep A555 (Abcam, ab150177), anti-mouse A488 (ThermoFischer Scientific, A-21202), anti-goat A488 (ThermoFischer Scientific, A-11055) and anti-mouse A555 (ThermoFischer Scientific, A-31570) were used as secondary antibodies. Images were acquired on an Axio Imager Z.2 (Zeiss) using the Imager Z.2 software, and analyzed with the Icy open source platform (55).

### Plasma cytokine quantification

IFN-γ and IL-17A concentrations in plasma were quantified by a high-sensitivity Simoa multiplex assay developed with Quanterix Homebrew kits, as previously described (56).

### 16S rRNA sequencing and analysis

The V3-V4 region of bacterial 16S rDNA was PCR amplified with V3-340F (CCTACGGRAGGCAGCAG) and V4-805R (GGACTACHVGGGTWTCTAAT) barcoded primers. PCR products were cleaned using Ampure magnetic purification beads (Agencourt AMPure XP Kit), quantified with the QuantiFluor® ONE dsDNA kit (Promega) and pooled in equal amounts of each PCR product. Library pools were loaded at 12pM with a 15% PhiX spike for diversity and sequencing control onto a v3 300-bp paired end reads cartridge for sequencing on the Illumina MiSeq NGS platform. After removing reads containing incorrect primer or barcode sequences and sequences with more than one ambiguous base, a total of 2.905.540 reads (121.064 mapped reads on average) was recovered from the 24 studied samples. Bioinformatics analysis was performed as described in (57).

### Statistics

The Graphpad Prism software (6.0; La Jolla, CA) was used for statistical comparisons and graphical representations. The statistical tests used are detailed in figure legends. Comparisons of metabolite levels were performed by linear model regression using the limma package in R software (53) and comparisons between lesions and matched healthy skin controls (L-HS vs H-HS) use a paired model, while comparisons between patients and healthy controls (HC) were corrected for gender and age difference by including those factors as confounders in the model. A false discovery rate (FDR) was calculated using the Benjamini-Hochberg method to account for multiple comparisons in analyses of metabolomic and imaging data. Differences corresponding to *FDR*<0.05 were considered significant.

### Study approval

This biomedical research study was approved by the French Ethics Committee (Comité de Protection des Personnes), the competent health authority (Agence Nationale de Sécurité des Médicaments et des produits de santé) and the French Data Protection Agency (Commission Nationale de l’Informatique et des Libertés). Study participants were identified by number. They provided written informed consent to the research, including data and biospecimen collection such as cutaneous biopsy sampling, prior to inclusion in the study.

## Data Availability

The data supporting the findings of this study are available within the article and supplementary materials.

## Author contributions

C.D., J.P.D., A.N. and M.D. conceived and designed the study. C.D., L.G.-M., J.-D.M. and J.-M.D. analyzed the data and C.D. wrote the manuscript. S.D. and A.H. provided primary fibroblasts and skin samples from HS patients and controls. M.D., A.N. and M.-N.U. conducted the recruitment of patients and healthy volunteers, and the collection of human samples. J.P.D., J.-M.D. designed and conducted the FACS analyses of PBMCs with assistance of L.G.-M. L.G.-M., J.-D.M., and V.M. designed and conducted all experiments using fibroblasts. L.G.-M. designed and conducted skin immunostaining studies with assistance of A.S. J.-D.M. analyzed metabolomics data. L.B. conducted the HPLC analysis of Trp metabolites in plasmas. D.D. supervised the SIMOA analysis of plasma cytokines performed by V.B. and P.G. designed, conducted the experiments and analysis of 16S rRNA sequencing.

## Acknowledgments

We thank Armanda Casrouge for assistance in performing the immunostaining of skin biopsies, Olivier Join-Lambert for fruitful discussions on the skin microbiota of HS patients and Harry Sokol for his critical reading of this manuscript. We also thank Valérie Briolat and Laurence Motreff of the Biomics Wet-Lab Platform for 16S rRNA sequencing, and the Image Analysis Hub for immunofluorescence data processing. This study was supported by Institut Pasteur (C.D., J.P.D.) via the Center for Translational Science (CRT), INSERM (U1221, C.D.; U1223, J.P.D.) and ‘Société Française de Dermatologie’. The Biomics Platform, C2RT, Institut Pasteur, Paris, France, is supported by France Génomique (ANR-10-INBS-09-09) and IBISA.

---

**Table S1.** Characteristics of study participants. (**A**) Metabolomic analysis of skin biopsies, assays of Trp metabolites and cytokines in plasma. (**B**) Transcriptomic analysis of skin biopsies and microbiota; *In 2 of the 8 HS patients, only H-HS skin samples were taken. (**C**) Primary fibroblasts.

|  | Healthy controls | Patients with HS |
| --- | --- | --- |
| **Total** | 20 | 19 |
| **Female, No (%)** | 12 (60) | 16 (84) |
| **Age, mean (SD)** | 46 (12) | 32 (6) |
| **Puncture site** | Axilla | Axilla (5), Groin (8), Buttock (6) |

**B**
|  | Healthy controls | Patients with HS |
| --- | --- | --- |
| **Total** | 9 | 8* |
| **Female, No (%)** | 8 (89) | 8 (100) |
| **Age, mean (SD)** | 34 (7) | 30 (8) |
| **Puncture site** | Abdomen (5), Leg (4) | Groin (4), Buttock (4) |

|  | Healthy controls | Patients with HS |
| --- | --- | --- |
| **Total** | 5 | 5 |
| **Female, No (%)** | 5 (100) | 5 (100) |
| **Age, mean (SD)** | 30 (8) | 30 (8) |
| **Puncture site** | Abdomen (3), Leg (1), Breast (1) | Axilla (1), Groin (2), Buttock (2) |

Table S2: Relative levels of metabolites in skin samples from healthy individuals and patients with Hidradenitis Suppurativa https://data.mendeley.com/datasets/4dk2x22ms6/draft?a=12f4c7fe-a2b3-403e-bc93-97c8f994b980

**Table S3.**
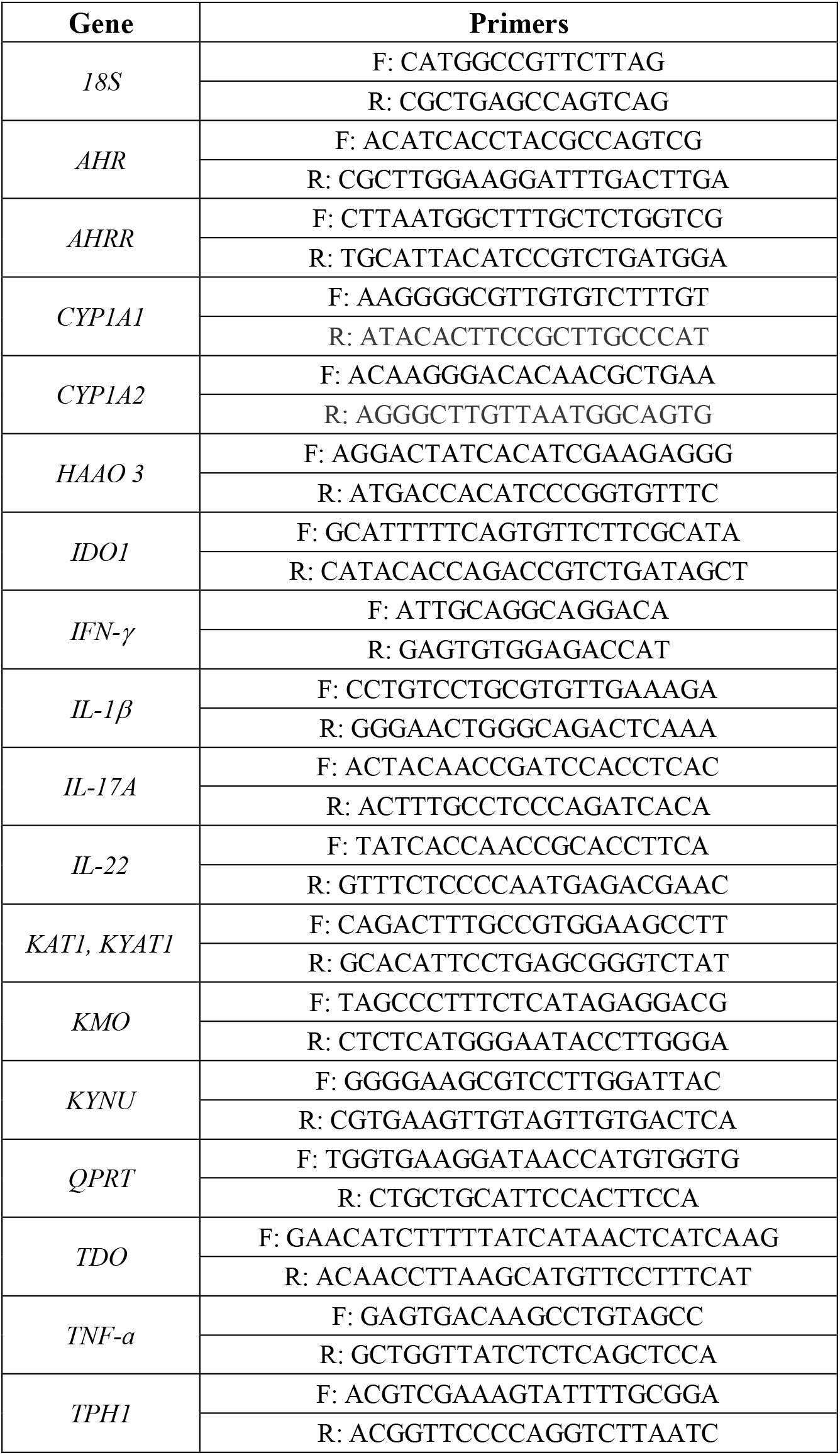
Primers used for qRT-PCR.

**Figure S1.**
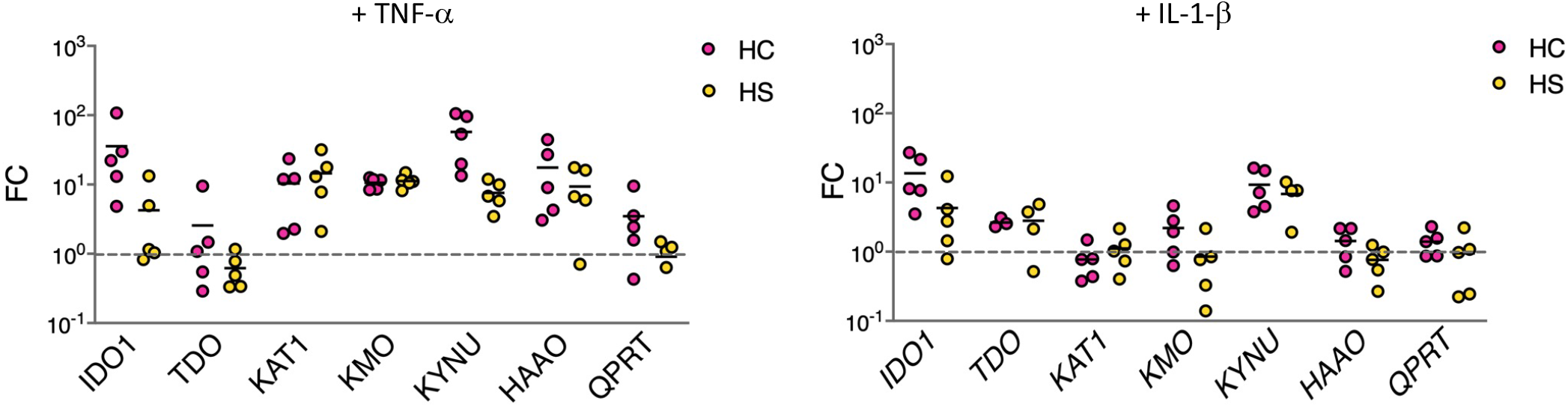
HS fibroblasts respond normally to *in vitro* stimulation with inflammatory cytokines. Fold change (FC) in expression of kynurenine pathway enzyme genes by primary fibroblasts from 5 HC and 5 HS patients following a 24h treatment with TNF-α (100 ng/ml) or IL-1β (2.5 ng/ml), relative to unstimulated controls. Data are shown as scatter dot plots with means.

**Figure S2.**
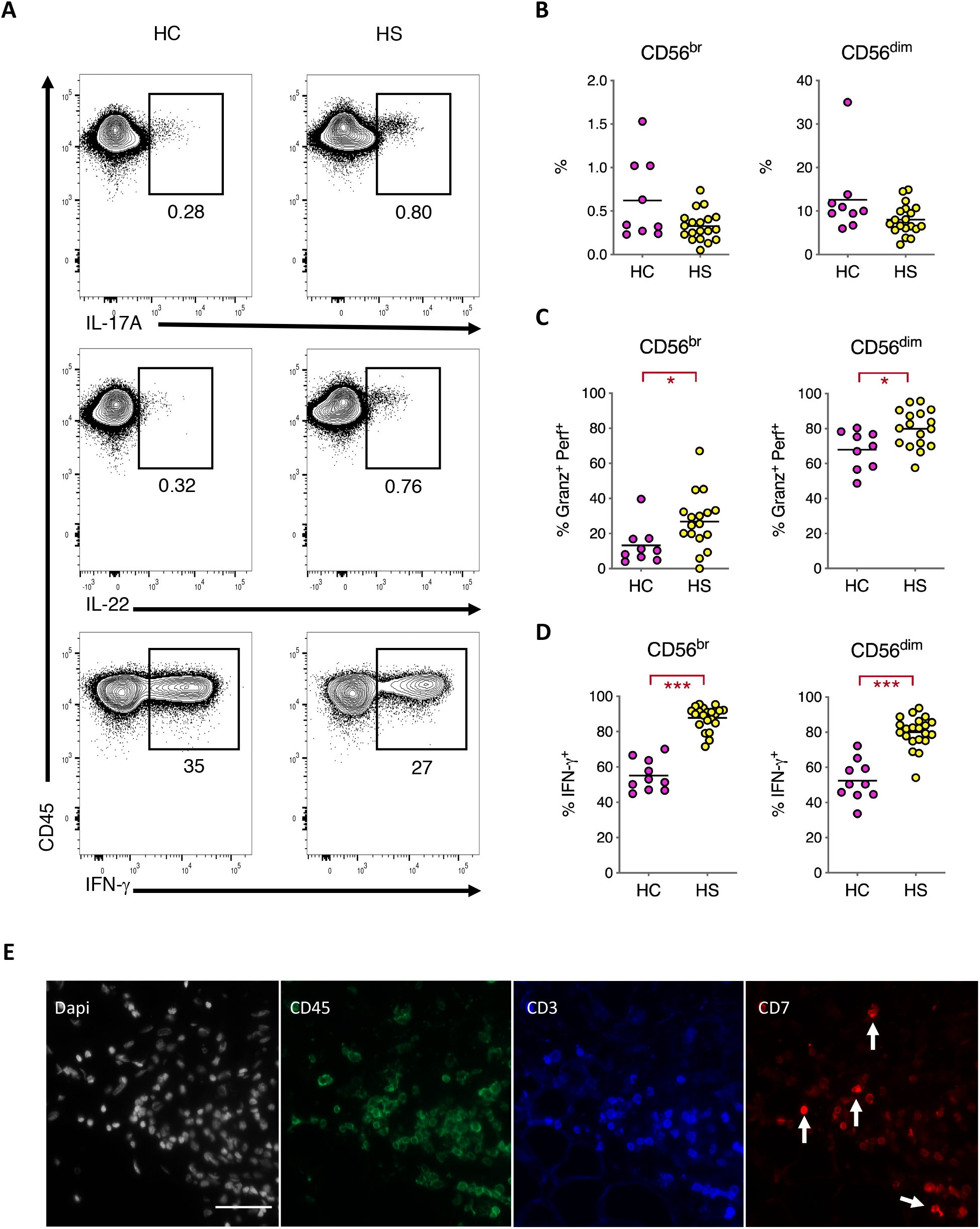
Immunological parameters differing between HS patients and HC controls. **(A)** Representative contour plots with percentages of IL-17A, IL-22 and IFN-g producing T cells (gated as CD3+ CD5+ CD45+) in HS patients and HC controls, following a 3h stimulation with PMA/Ionomycin. 100,000 event per contour plots are depicted for comparison. (**B**) Frequency of CD56br and CD56dim NK cells, amongst total lymphocytes, in HC and HS patients. (**C**) Proportion of CD56br and CD56dim NK cells co-expressing Granzyme B and Perforin. (**D**) Proportion of CD56br and CD56dim NK cells producing IFN-γ following a 3h stimulation with PMA/Ionomycin. Data are shown as scatter dot plots with means. *P<0.05, ***P<0.001 by Mann-Whitney U-test. (**E**) DAPI, CD45, CD3 and CD7 staining of a representative L-HS skin section, with arrows pointing to CD3^-^ CD7+ NK cells. Scale bar 50 μm.

**Figure S3.**
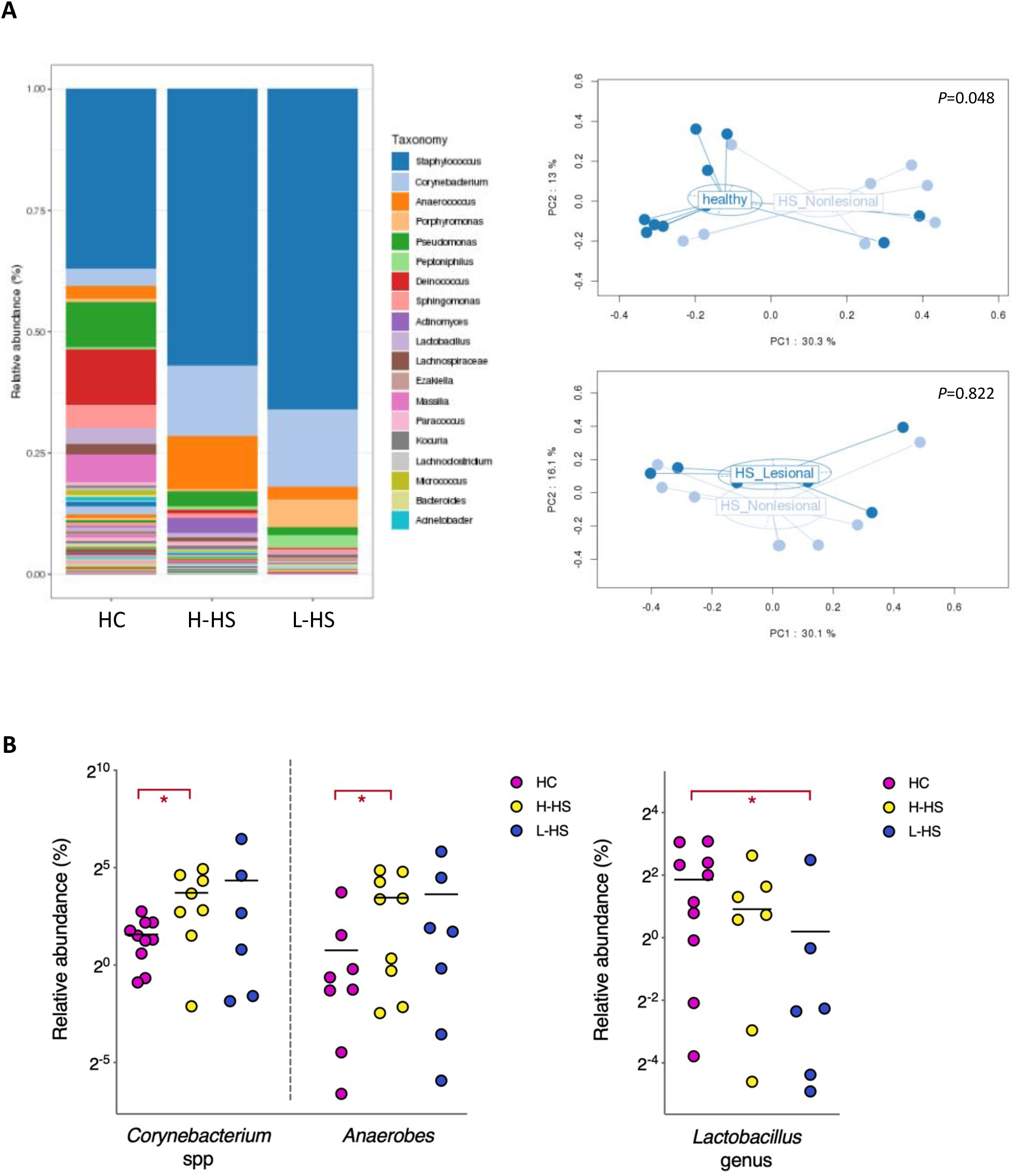
Changes in the bacterial composition of healthy and lesional skin microbiomes. (**A**) Genus distribution and PCA analysis of the bacterial species found in HC, H-HS and L-HS skin biopsies. *P* values by Permanova test. (**B**) Relative abundance of *Corynebacterium* spp, pooled anaerobes (*Prevotella*, *Porphyromonas*, *Actinomyces* and *Anaerococcus* spp) and *Lactobacillus* spp in HC, H-HS and L-HS skin samples. \**P*<0.05, by Mann-Whitney U-test.

## Notes

### Competing Interest Statement

The authors have declared no competing interest.

### Author Declarations

This biomedical research study was approved by the French Ethics Committee, the competent health authority and the French Data Protection Agency. Study participants were identified by number. They provided written informed consent to the research, including data and biospecimen collection such as cutaneous biopsy sampling, prior to inclusion in the study.

